# Bridging Specialist Gaps: Outcomes and Lessons from a Surgical Outreach Initiative in Rural Ghana as a Pathway to Universal Health Coverage

**DOI:** 10.1101/2025.08.01.25332723

**Authors:** Samuel Mayeden, Amponsah Foster, Arko Akoto-Ampaw, Temitope Ebenezer Arkorful, Frank Ewusi-Brown, Ziblim Adam Kasule, Gasinu Joseph, Adbul Malik Tanko, Isaac Bentum Ogoe, James Mawusi Adzraku, Peter Dambach, Andreas Deckert, Micheal Marx, Justice Yevugah Sitsofe, John Koku Awoonor-Williams

## Abstract

**Background:** Limited access to essential surgical care in rural Ghana contributes to health inequities and hinders progress toward Universal Health Coverage. Integrating surgical services into primary health care is vital in resource-limited settings.

**Methods:** We implemented a specialist-led surgical outreach in Kwahu Afram Plains North District, Ghana, using community mobilization and interdisciplinary collaboration. Patients were screened for medical eligibility and NHIS coverage. Data on demographics, diagnoses, procedures, outcomes at 14, 30, and 90 days, and programme costs were collected and analyzed descriptively.

**Results:** Of 200 patients screened, 185 were eligible for surgery. Inguinal hernia, uterine fibroids, and non-toxic goitre were the most common conditions. All patients achieved successful post-operative outcomes at each time point. Programme costs totaled 216,000 Ghana cedis, with NHIS and civil society support.

**Conclusion:** Specialist-led surgical outreach, integrated into primary care, can effectively expand surgical access and support progress toward UHC in rural, resource-constrained settings.

## 1. Introduction

Universal Health Coverage (UHC) aims to ensure that all individuals receive the healthcare services they need without financial hardship (WHO, 2010). Integrating essential surgical care into Primary Health Care (PHC) is a critical but often underappreciated aspect of UHC, particularly in resource-limited settings (Funk et al., 2010). Surgical care, though frequently perceived as a higher-level service, is indispensable in addressing a wide range of health conditions, including trauma, congenital anomalies, non-communicable diseases, and obstetric complications (Mock et al., 2015).

In Low- and Middle-Income Countries (LMICs) such as Ghana, the burden of surgically treatable diseases is substantial. An estimated 28–32% of the global burden of disease can be addressed by surgery (Shrime et al., 2015), yet access to safe, timely, and affordable surgical care remains limited in many LMICs due to inadequate infrastructure, workforce shortages, and financial barriers (Kruk et al., 2018). These challenges are especially pronounced in rural and underserved areas, where approximately 60% of Ghana’s population resides and where health facilities often lack even basic surgical capabilities (Osen et al., 2011)

Surgical outreach programmes have become a vital strategy for addressing these gaps. By sending teams of healthcare professionals to rural settings, these programmes both provide direct surgical services and build local capacity through training and mentorship (Galukande et al., 2010) Evidence from Ghana and similar settings shows that such initiatives can improve access to essential surgical care, increase the number of surgeries performed, and enhance the skills of local healthcare workers, contributing to a more sustainable health system (Ameh et al., 2012).

Integrating surgical care into PHC is essential for achieving UHC. PHC is the first point of contact for most patients and plays a crucial role in early diagnosis and management of surgical conditions (Farmer & Kim, 2008). Comprehensive care that includes surgical services at the PHC level can address the full spectrum of health needs, from prevention to rehabilitation. Achieving this integration requires investments in health infrastructure, workforce development, and sustainable financing mechanisms (Funk et al., 2010).

Despite Ghana’s progress in many health indicators, substantial challenges remain in ensuring equitable access to surgical care. Disparities between urban and rural areas persist, with rural populations facing limited access to specialist surgeons, anesthetists, and diagnostic tools (Anyfantakis et al., 2012). Innovative solutions such as surgical outreach programmes and the integration of surgical care into PHC are critical to addressing these inequities and achieving national UHC goals (Groen et al., 2015; Kruk et al., 2018).

The Kwahu Afram Plains North District exemplifies these challenges. The district’s only hospital operates with a severely limited staff, just two medical officers, two certified registered anesthetists, and one critical care nurse, supported by a small auxiliary team. The lack of essential diagnostic equipment, such as an X-ray machine, further hampers effective care, forcing clinicians to rely on presumptive diagnoses. These constraints result in delayed or unmet surgical care, particularly for complex cases, and often compel patients to travel long distances at considerable personal and financial cost (Osen et al., 2011)

Prior to the surgical outreach described in this study, many indigent patients in the district remained untreated or received suboptimal care. The outreach programme was developed as a direct response to this gap, aimed at providing specialist-led surgical services and building local capacity within the district hospital. This article describes the implementation of the outreach using an interdisciplinary approach to patient screening, surgical intervention, and post-operative care, and details the strategies employed to overcome resource limitations and specialist shortages, with the aim of improving healthcare delivery across rural districts in Ghana.

Figure 1 shows the location of Kwahu Afram Plains North District in the Eastern Region of Ghana, the site of the surgical outreach initiative

**Figure 1.**
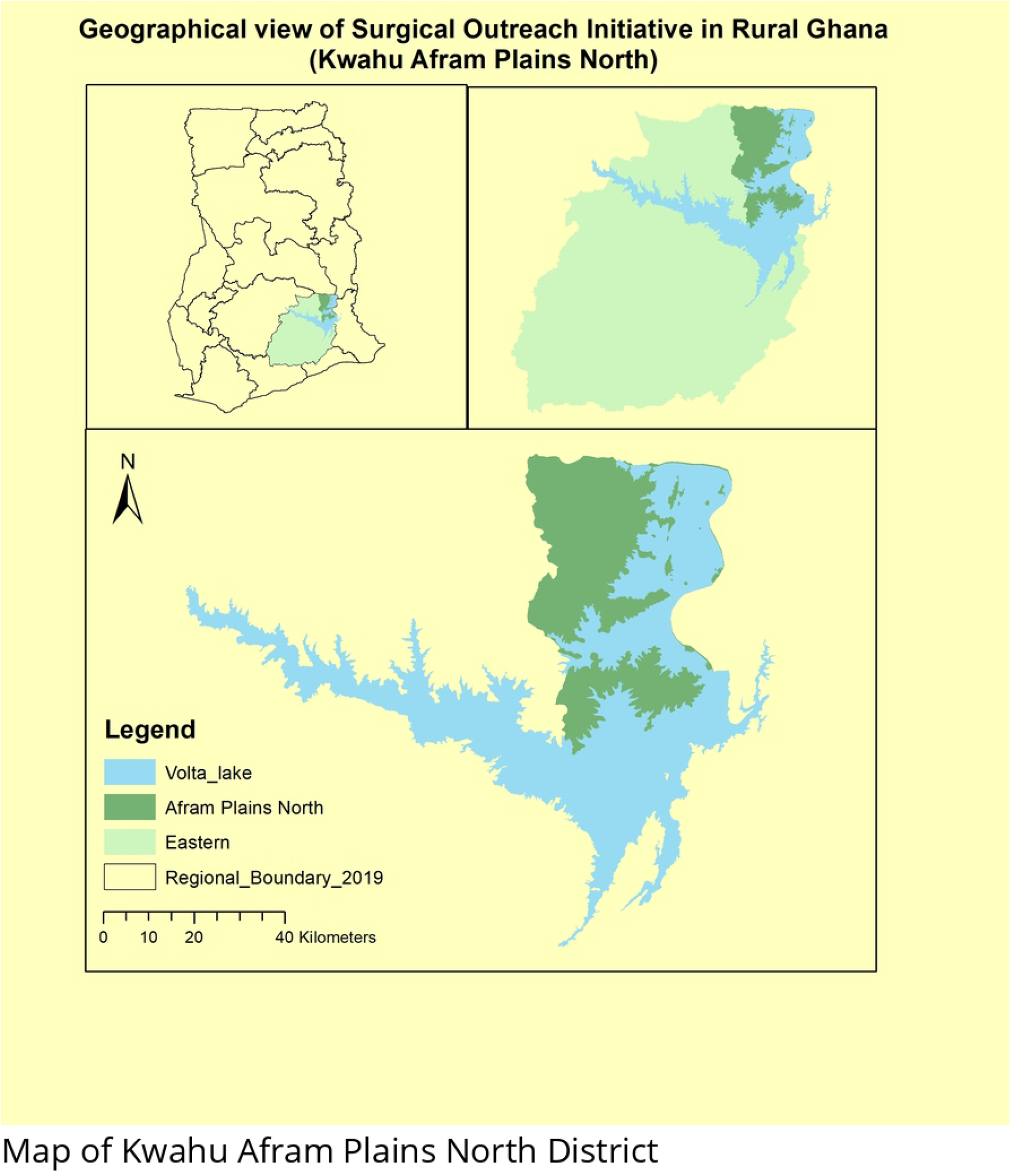
Map of Kwahu Afram Plains North District in Ghana. The location of the district is shown within the Eastern Region of Ghana, highlighting the catchment area for the surgical outreach programme.

## 2. Methodology

We implemented a specialist-led surgical outreach intervention in the Kwahu Afram Plains North District to address critical gaps in access to essential surgical care (Mayeden et al., 2023). This initiative was developed through collaboration among Donkorkrom Presbyterian Hospital, the Kwahu Afram Plains North District Assembly, the Career Guidance and Mentorship Foundation, and the Eastern Regional Hospital, Koforidua. Each partner contributed resources and expertise to meet the needs of the district’s underserved population (Asare et al., 2021)

To maximize community engagement, we launched a broad publicity campaign using local radio stations, community information centers, and posters to raise awareness about the outreach and the process for patient enrollment (Owusu-Ansah et al., 2020).

Eligibility criteria were established to prioritize urgent surgical needs and ensure efficient use of limited resources (Ameh et al., 2012). Potential candidates were required to demonstrate medical fitness and have a condition aligned with the surgical services offered. Possession of a valid National Health Insurance Scheme (NHIS) card was mandatory to facilitate partial reimbursement to the hospital (National Health Insurance Authority, 2019).

Patient screening was conducted over five days (December 2–7, 2022). Healthcare professionals performed comprehensive assessments to determine the suitability of each patient for surgery, ensuring timely and equitable access to care for those most in need.

### 2.1 Ethical Considerations

This surgical outreach was conducted as a health system strengthening and quality improvement activity under the routine operations of Donkorkrom Presbyterian Hospital. According to hospital policy and local regulations, formal ethical approval was not required for this programmatic work. All patient data were anonymized before analysis, and no personal identifiable information was collected or reported. Verbal consent was obtained from all participants or their legal guardians prior to surgical care and inclusion in the analysis (Gyamfi et al., 2022).

## 3. Key Findings

### 3.1 Age Distributions and Baseline Characteristics

Table 1 summarizes the age and gender characteristics of patients treated during surgical outreach. Of the 200 patients screened, 185 were eligible for surgery and 15 were excluded based on clinical criteria. The mean age was 44 years (range: 1–83 years), and the cohort consisted of 54% males and 46% females. All eligible patients possessed valid NHIS cards. The most common diagnoses were inguinal hernia, uterine fibroids, and non-toxic goitre. The most frequently performed procedures included hernia repair, myomectomy, and thyroidectomy.

**Table 1:** Baseline Characteristics of Patients Eligible for Surgery.

| Characteristic | Value |
| --- | --- |
| Total Patients Screened | 200 |
| Eligible Patients | 185 |
| Excluded Patients | 15 |
| Mean Age (years) | 44 |
| Age Range (years) | 1–83 |
| Male (%) | 54 |
| Female (%) | 46 |
| NHIS Card Holders (%) | 100 |
| Common Diagnoses | Inguinal Hernia, Uterine Fibroids, Non-toxic Goitre |
| Most Frequent Procedures | Hernia Repair, Myomectomy, Thyroidectomy |

The outreach began with community mobilization, patient sensitization, and clinical screening. Individuals meeting the eligibility criteria were scheduled for surgery, while others received counseling and referral for appropriate follow-up care. These findings highlight the breadth of surgical needs in the community and underscore the critical role of outreach programmes in addressing them.

### 3.2 Frequency of Diagnosis Among Patients

Figure 2 presents the frequency of diagnoses among eligible patients. Inguinal hernia was the most prevalent condition, diagnosed in 82 patients (44%). Uterine fibroids were identified in 36 patients (19%), and non-toxic goitre in 25 patients (14%). Other less common conditions included hydrocele (10 cases, 5%) and lipoma (8 cases, 4%). These data provide a clear snapshot of the surgical health needs within the district and highlight the substantial demand for specialized surgical services, particularly for hernia repair, myomectomy, and thyroidectomy.

**Figure 2.**
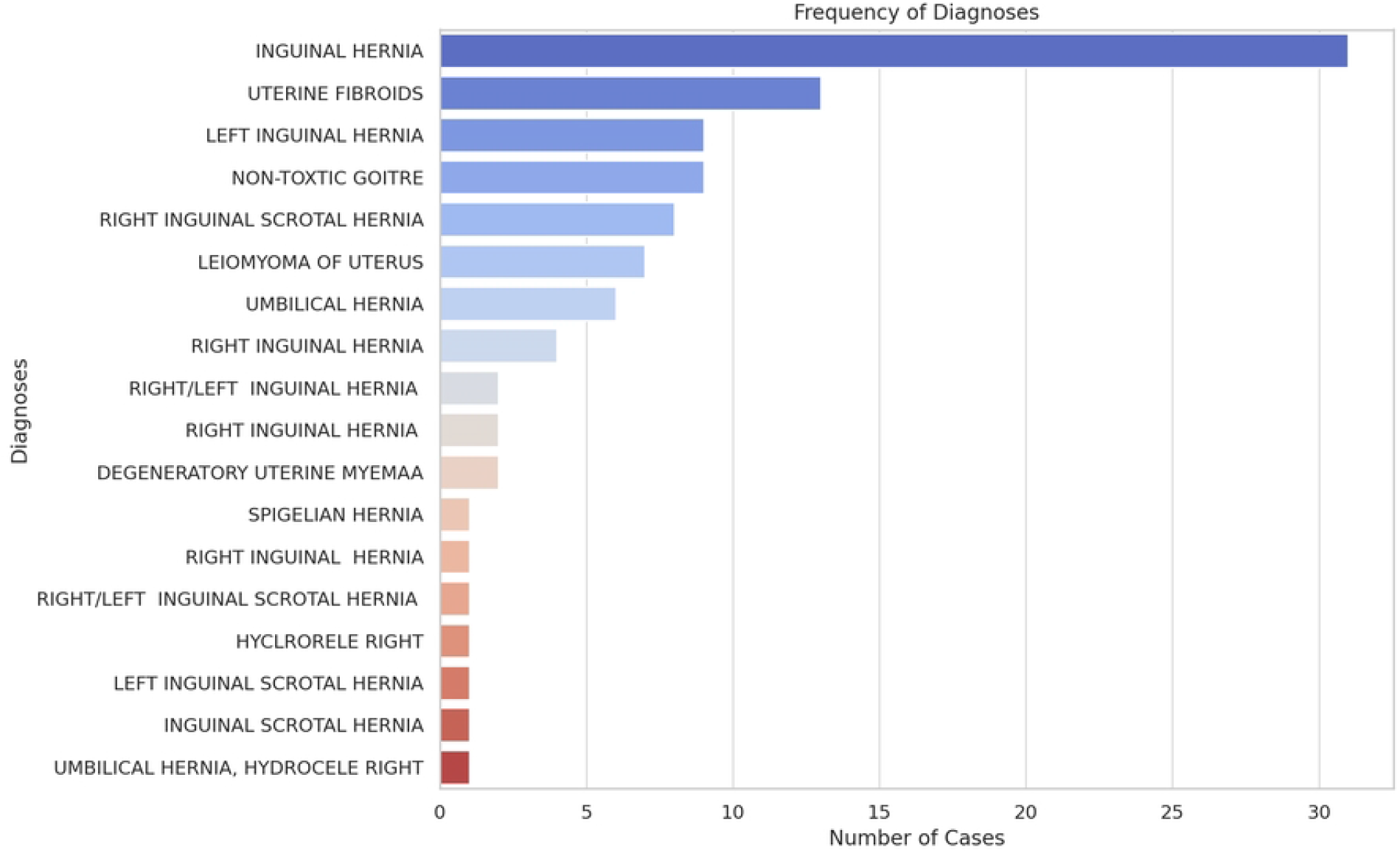
Distribution of Surgical Diagnoses Among Eligible Patients. The frequency of key diagnoses among 185 patients eligible for surgery. Inguinal hernia was the most common diagnosis (44%), followed by uterine fibroids (19%), non-toxic goitre (14%), hydrocele (5%), and lipoma (4%).

Figure 2 illustrates the frequency of major surgical diagnoses identified among patients eligible for the outreach, highlighting the predominance of inguinal hernia, uterine fibroids, and non-toxic goitre.

### 3.3 Post-Operative Outcomes

Figure 3 summarizes the post-operative outcomes, with follow-up assessments conducted at 14, 30, and 90 days after surgery. Success was defined as a clean surgical site at 14 days, clinical stability at 30 days, and evidence of improvement at 90 days. All patients met these criteria, resulting in a 100% success rate: all exhibited a clean surgical site at day 14, remained stable on day 30, and showed improvement by the 90-day follow-up.

**Figure 3.**
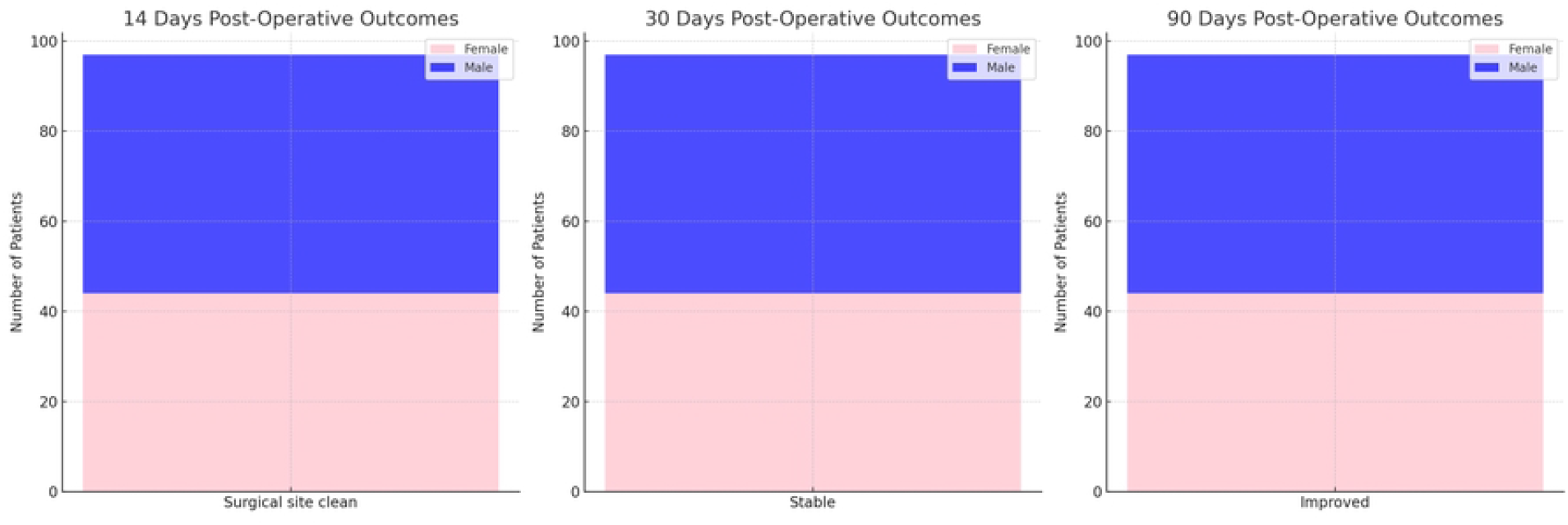
Post-Operative Outcomes at 14, 30, and 90 Days. Proportion of patients achieving defined post-operative success at follow-up intervals: clean surgical site at 14 days, clinical stability at 30 days, and improvement at 90 days. All patients met the success criteria at each time point.

Further analysis by sex showed no significant differences in recovery rates; both male and female patients experienced consistent and successful outcomes across all measured intervals. These results confirm the effectiveness of the surgical procedures performed and suggest positive outcomes regardless of patient demographics.

Post-operative outcomes at 14, 30, and 90 days following surgery are summarized in Figure 3, demonstrating the uniformly positive recovery rates across all time points.

### 3.4 Operational Costs

Figure 4 outlines the financial aspects of surgical outreach. A total of 216,000 Ghana cedis (approximately $22,736.84) was allocated to support patient care, covering both surgical procedures and medications. The average cost per patient was 1,500 Ghana cedis ($157.89), which included costs covered by NHIS and required top-up payments. The NHIS played a crucial role in ensuring access to care without undue financial burden for patients’ families.

**Figure 4.**
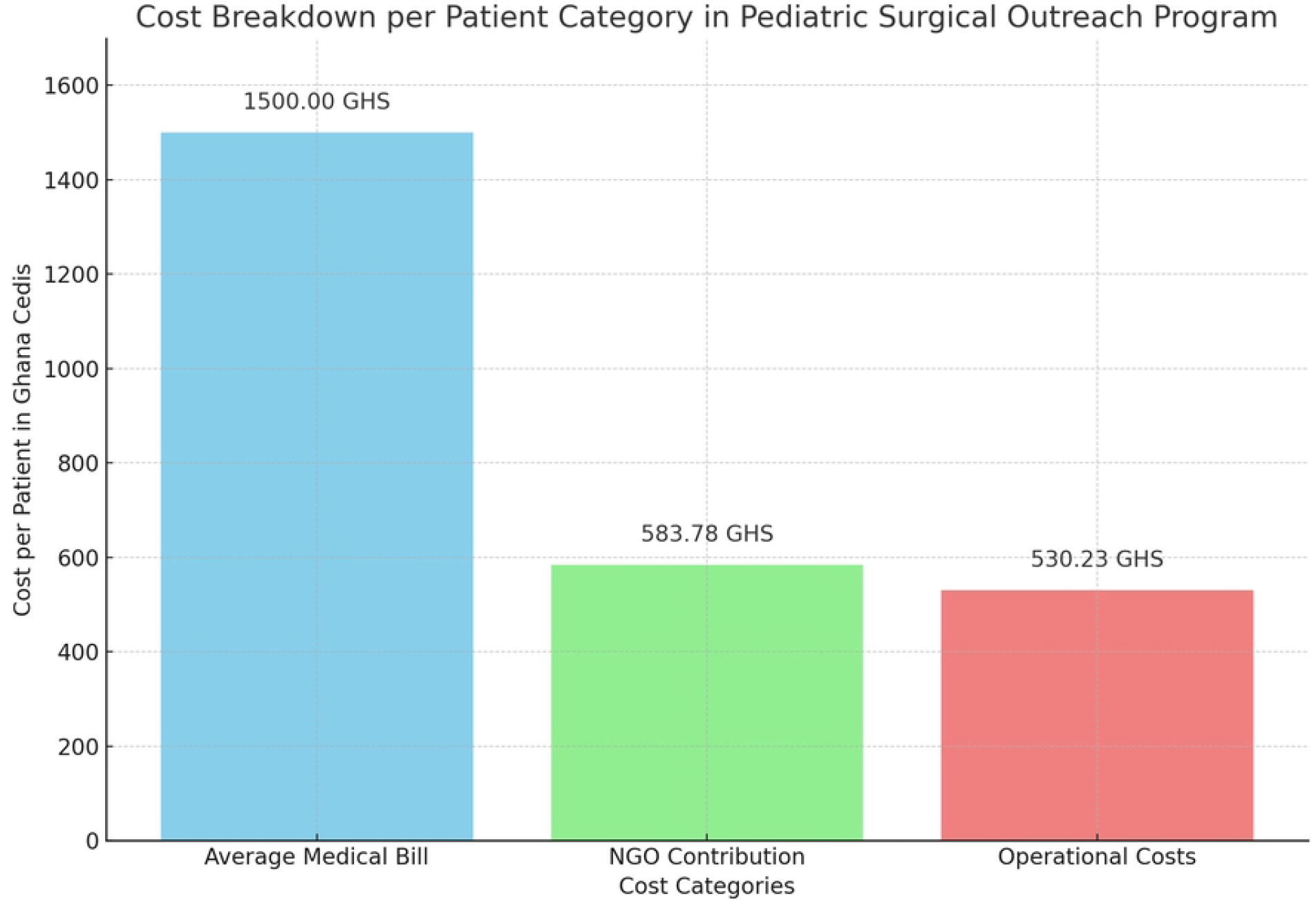
Breakdown of Programme Operational Costs. Allocation of total outreach expenditures (216,000 Ghana cedis, ∼$22,736.84 USD) by category, including direct surgical care, medications, and operational/logistics costs, with contributions from NHIS and civil society organizations.

Half of the total cost (108,000 Ghana cedis; $11,368.42) was provided by the Civic Society Organization (CSO), which significantly offset expenses and enabled care for a larger number of patients. In addition to direct medical costs, operational expenses—such as per diem, accommodation, feeding, and transportation for clinicians—totaled 98,092 Ghana cedis ($10,325.47). These operational costs were essential for deploying skilled professionals to remote areas and expanding access to specialized surgical care.

Figure 4 presents a breakdown of the operational costs associated with the surgical outreach programme, including patient care, medications, and logistics.

## 4. Discussions

Our study provides valuable insights into the demographics, diagnoses, and outcomes of a specialist-led surgical outreach programme in the Kwahu Afram Plains North District. The intervention successfully treated 185 patients, with a mean age of 44 years and a balanced gender distribution (54% male, 46% female).

Our analysis of age and gender distributions revealed a balanced median age for both groups, indicating the need for surgical interventions across the entire age spectrum. Males exhibited greater age variability, potentially reflecting different health-seeking behaviors or a broader range of surgical conditions. The presence of outliers in both gender groups emphasizes the programme’s capacity to serve a diverse population (Alkire et al., 2015; Funk et al., 2010)

Our findings show that inguinal hernia emerged as the most common diagnosis, underscoring a substantial need for surgical care in this rural community. This high prevalence may be linked to the occupational profile of the district, where many residents engage in strenuous physical labor, a known risk factor for hernias (Ameh et al., 2012; Groen et al., 2015). Other prevalent conditions, such as uterine fibroids and non-toxic goitre, highlight ongoing health challenges, particularly among women. These findings point to a persistent need for specialized surgical services in the Afram Plains region and demonstrate the outreach programme’s critical role in addressing these needs (Kruk et al., 2018; Mock et al., 2015; Osen et al., 2011)

Our post-operative outcomes demonstrate a high level of success, with all patients meeting the defined success criteria at 14, 30, and 90 days after surgery. Both male and female patients experienced consistent positive outcomes, underscoring the high quality and reliability of surgical care delivered during the outreach. These results support the effectiveness of the programme in providing successful interventions regardless of patient demographics (Funk et al., 2010).

Our financial analysis highlights the significant resources required to provide comprehensive surgical care in resource-limited settings. The programme’s total expenditure of 216,000 Ghana cedis ($22,736.84), including surgical procedures and medications, underscores the financial commitment needed for sustainability. The NHIS played a pivotal role in covering a substantial portion of patient costs, and the local NGO’s contribution of 108,000 Ghana cedis ($11,368.42) significantly reduced the burden on patients. Additional operational costs for logistics and accommodation (98,092 Ghana cedis; $10,325.47) further illustrate the investment required to deploy skilled clinicians to remote areas and expand access to care (Funk et al., 2010; Gyamfi et al., 2022; Kruk et al., 2018; Mock et al., 2015)

## 5. Lessons Learned

Our experience with the surgical outreach initiative in the Afram Plains district revealed several important lessons for expanding access to surgical care and progressing toward universal health coverage in resource-limited settings (Ameh et al., 2012). First, we found that strong interdisciplinary collaboration among hospitals, local authorities, regional specialists, and civil society organizations was critical for mobilizing the expertise and logistical support needed to deliver services that no single institution could have achieved alone (Funk et al., 2010). Community engagement, especially through local radio, volunteers, and district assemblies, played a central role in building awareness, trust, and participation across diverse demographic groups (Groen et al., 2015).

We also learned that while requiring valid NHIS cards enabled cost recovery for the hospital and supported programme feasibility, it inadvertently excluded some uninsured or informally registered individuals. This highlights the need for broader health insurance enrollment and more flexible financing mechanisms for marginalized populations (National Health Insurance Authority, 2019). The shortage of local specialists and diagnostic infrastructure made us dependent on external teams and equipment; although this addressed immediate needs, it underscored the importance of long-term investment in local workforce development, equipment maintenance, and task-sharing to ensure continuity when external partners are not present.

Despite the immediate impact of outreach, sustainability remains a significant challenge. Reliable funding, integration into routine health services, and government support are essential for transforming such interventions from ad hoc efforts into institutionalized components of primary healthcare (Mock et al., 2015). Ongoing evaluation and adaptive management were valuable for quickly identifying service gaps and logistical bottlenecks, suggesting that future programmes would benefit from more robust data systems and clear indicators of success, such as patient outcomes, cost-effectiveness, and equity in service delivery (Kruk et al., 2018).

Ultimately, the outreach model we implemented demonstrates that interdisciplinary collaboration, community mobilization, and partial cost recovery through health insurance can be effective strategies in similar contexts (Galukande et al., 2010). However, each setting requires adaptation to address local realities, including risks of exclusion and challenges to sustainability.

## 6. Challenges and Strategic Recommendations

Delivering sustainable surgical care in rural, resource-limited settings such as the Afram Plains District presents significant challenges, but also clear opportunities for strategic improvement (Mock et al., 2015). One of the primary issues is the sustainability of outreach initiatives. While short-term surgical missions are effective in addressing immediate health needs, they do not offer a lasting solution to the persistent gaps in rural surgical care, and reliance on these periodic activities underscores the absence of enduring health system capacity in many rural areas of Ghana (Ameh et al., 2012).

Weak healthcare infrastructure and a limited workforce further complicate the situation. The lack of essential diagnostic tools, like X-ray machines, and shortages of specialized personnel such as surgeons and anesthetists make the delivery of safe and timely surgical care difficult (Osen et al., 2011). The geographic isolation of the Afram Plains compounds these barriers, making it challenge both to attract and retain healthcare professionals and for patients to access care, especially in emergencies (Funk et al., 2010). Financial constraints continue to be a concern for both patients and outreach programmes. Despite support from philanthropists and the NHIS, patients may still face out-of-pocket costs, and the organization of outreach activities remains resource-intensive, requiring significant funding for logistics, accommodation, and medical supplies (National Health Insurance Authority, 2019).

To overcome these challenges and facilitate the integration of essential surgical services into primary health care, several strategic actions are needed. Investing in local capacity building is essential ongoing training for local healthcare providers and the establishment of regional mentorship networks can build long-term competencies and gradually reduce dependency on visiting teams. Upgrading hospital infrastructure and equipment, particularly operating theaters, diagnostic tools, and recovery spaces in rural district hospitals, should be a priority for government and development partners. Strengthening incentives to attract and retain skilled health workers such as offering housing, professional development, and career advancement opportunities can help address workforce shortages (Groen et al., 2015).

Improving financing models is equally important. Expanding NHIS coverage for surgical services and leveraging partnerships with NGOs, local governments, and donors can help reduce hidden costs for patients and support programme sustainability (Kruk et al., 2018). Additionally, developing coordinated referral systems and providing support for patient transport are crucial to ensuring that individuals in remote areas can access timely surgical care when needed (Alkire et al., 2015). Together, these strategic investments and policy actions can address current challenges and create a more resilient, equitable, and sustainable model for surgical care in rural Ghana.

## 7. Policy Implications and Conclusion

This surgical outreach programme demonstrates that essential surgical care can be delivered effectively in resource-limited rural settings through interdisciplinary collaboration. However, ensuring long-term sustainability and equity requires strategic policy actions. Policymakers should prioritize integrating surgical services into primary health care by investing in rural infrastructure, expanding NHIS coverage for surgical conditions, and building the capacity of local health workers. Strong partnerships among government, civil society, and local health systems are vital for mobilizing resources and sustaining outreach activities. Embedding such models within national UHC strategies can help close the surgical access gap and improve health equity.

Our experience in Afram Plains district shows that targeted outreach can provide timely, effective surgical care for populations with limited access, yielding uniformly positive outcomes. Integrating surgical services into primary care is both feasible and valuable in advancing Universal Health Coverage. To sustain and expand such programmes, ongoing investment in infrastructure, workforce, and financing is essential. Scaling and institutionalizing outreach models within district health systems will reduce inequities, strengthen system resilience, and accelerate progress toward national and global health goals.

## 8. Data Availability Statement

The data supporting the findings of this study are available from the corresponding author upon reasonable request. Due to privacy and ethical considerations related to patient health records, some data may not be publicly shared.

## 9. Ethics Statement

This initiative was conducted as a health system strengthening activity and did not require formal ethical approval, as per local guidelines. Consent was obtained from all participants or their guardians.

## 12. Acknowledgments

The authors gratefully acknowledge the dedication of the clinical and administrative staff of Donkorkrom Presbyterian Hospital, the Kwahu Afram Plains North District Assembly, and the supporting civil society organization. Special gratitude goes to the ERHK, whose specialist team provided invaluable expertise and support during the outreach program. We especially thank the volunteers and local radio stations that contributed to community mobilization efforts. Special appreciation also goes to the patients and families who participated in the outreach programme.

